# Public perceptions of COVID-19 in Australia: perceived risk, knowledge, health-protective behaviours, and vaccine intentions

**DOI:** 10.1101/2020.04.25.20079996

**Authors:** Kate Faasse, Jill Newby

## Abstract

Widespread and sustained engagement with health-protective behaviours (i.e., hygiene and distancing) is critical to successfully managing the COVID-19 pandemic. Evidence from previous emerging infectious disease outbreaks points to the role of perceived risk, worry, media coverage, and knowledge in shaping engagement with health-protective behaviours as well as vaccination intentions. The current study examined these factors in 2,174 Australian residents. An online survey was completed between 2-9 March 2020, at an early stage of the COVID-19 outbreak in Australia. Results revealed that two thirds of respondents were at least moderately worried about a widespread COVID-19 outbreak in Australia (which subsequently occurred). Worry about the outbreak and closely following media coverage were consistent predictors of health-protective behaviours (both over the previous month, and intended behaviours in the case of a widespread outbreak) as well as vaccination intentions. Health-behaviour engagement over the previous month was lower in some demographic groups, including males and younger individuals (18-29 age group). These was a substantial mismatch between respondents’ expected symptoms of infection and emerging evidence that a meaningful proportion of people who contract the novel coronavirus will experience asymptomatic infection (i.e., they will not experience symptoms associated with COVID-19). Only 0.3% of those in the current study believed that they personally would not experience any symptoms if they were infected. Uncertainty and misconceptions about COVID-19 were common, including one third of respondents who reported being unsure whether people are likely have natural or existing immunity. There was also uncertainty around whether specific home remedies (e.g., vitamins, saline rinses) would offer protection, whether the virus could spread via the airborne route, and whether the virus was human made and deliberately released. Such misconceptions are likely to cause concern for members of the public. These results point to areas of uncertainty that could be usefully targeted by public education campaigns, as well as psychological and demographic factors associated with engagement with health-protective behaviours. These findings offer potential pathways for interventions to encourage health-protective behaviours to reduce the spread of COVID-19.

## Introduction

On December 31, 2019, the first report of a ‘pneumonia of unknown cause’ was made to the World Health Organization (WHO) Country Office (World Health Organization, 2020c). The report came from a case in Wuhan, China. On January 10, 2020, WHO issued its first guidance on the ‘novel coronavirus’, with similarities to other coronaviruses such as SARS (Severe Acute Respiratory Syndrome) and MERS (Middle East Respiratory Syndrome). By the end of January, the novel coronavirus had spread to countries around the world, and the outbreak was declared a Public Emergency of International Concern, with 7834 confirmed cases and 170 people who had died from the novel coronavirus (now named SARS-CoV-2, the virus which causes the coronavirus disease COVID-19). The first cases of COVID-19 in Australia were identified on January 25, in the states of Victoria and New South Wales (Minister for Health, 2020). On February 1, Australia announced travel restrictions for foreign nationals who had been in high risk regions, and required 14-day self-isolation for returning citizens and permanent residents (Prime Minister of Australia, 2020). As of March 7, 2020, there were 63 confirmed cases of COVID-19 in Australia, including 2 deaths, with the majority (28) of cases in New South Wales, followed by 11 in Queensland (Australian Government Department of Health, 2020b). Most of these cases have been linked to direct or indirect travel to regions with know COVID-19 outbreaks, however, 10 cases did not have reported travel history, and instead occurred following person-to-person transmission within Australia.

This rapidly evolving situation has generated substantial media coverage, uncertainty about the situation and the state of scientific knowledge about the virus, and public anxiety about the potential risks associated with COVID-19 (Asmundson & Taylor, 2020). Public perceptions of risk, such as the perceived likelihood (how likely a person thinks it is that they will become infected) and severity of infection (how severe the symptoms would be if they were infected), have been shown to predict the degree to which individuals engage in – or intend to engage in – protective behaviours during infectious disease outbreaks (Petrie, Faasse, & Thomas, 2016; Weinstein, 1988). For example, higher perceived likelihood and severity of influenza A/H1N1 (swine flu), influenza H5N1 (bird flu), and SARS were associated with increased heath-protective behaviours (e.g. handwashing) in general population samples (Lau, Kim, Tsui, & Griffiths, 2007; Rubin, Amlot, Page, & Wessely, 2009; Tang & Wong, 2003). In addition, health-protective behaviours during a pandemic are associated with more accurate knowledge about the virus (Petrie et al., 2016) and the perception that these behaviours will be effective in reducing the risk of infection, as well as heightened anxiety, and trust in information provided by authorities (Bish & Michie, 2010).

During past pandemics, worry and anxiety have typically been high, with between one fifth and one third of people feeling very or extremely worried about contracting viruses such as SARS (Bults et al., 2011; Wheaton, Abramowitz, Berman, Fabricant, & Olatunji, 2012). Anxiety and risk perceptions may be influenced by media coverage and exposure to misinformation about the emerging health threat of COVID-19. Even with accurate media reporting – a particular challenge in a changing situation in which scientific information about the virus is emerging and being disseminated in real-time – the amount, framing, tone, sources, and perceived trustworthiness of media coverage can affect public risk perceptions (Paek & Hove, 2017). Analysis of media coverage during the Influenza A/H1N1 (swine flu) pandemic shows a large volume of news stories that tended to present conflicting and threatening information, while neglecting coverage of self-protective behaviours (Klemm, Das, & Hartmann, 2016). Even subtly misleading information, for example through the use of inaccurate images or misleading headlines, can bias readers’ interpretation of and memory for the information (Ecker, Lewandowsky, Chang, & Pillai, 2014). In addition to a large volume of media coverage, misinformation and conspiracy theories regarding COVID-19 are widespread and still emerging and evolving at the time of writing (Wikipedia, 2020; World Health Organization, 2020a). Such misinformation can have lasting impacts, including reduced engagement with health-protective behaviours including vaccination, once it becomes available (Zimet, Rosberger, Fisher, Perez, & Stupiansky, 2013).

The current study investigated the Australian public’s perception of risk (i.e. likelihood and severity) and worry about COVID-19, as well as sources of information, accuracy of current knowledge (and conversely, misinformation) about the virus, and current and intended protective health behaviours. Insight into how perceptions of emerging infectious diseases influence both actual and intended adoption of health protective behaviours is important in understanding the potential health, social, and economic impact of such outbreaks, and may contribute to targeting public health messaging to encourage appropriate health behaviours.

## Materials and Methods

### Recruitment

Members of the Australian general population were recruited for the online survey by the use of Facebook advertisements targeted at all users with current country of residence listed as Australia, and age listed as 18 or above, as well as one Facebook post (to a university page). Advertising and data collection ran for seven days from 4pm Monday March 2, to 4pm Monday March 9, 2020. In total the ad was displayed to 66,210 individual accounts, with 4,353 clicks. Each response came from a unique IP address.

### Ethical approval

The study was approved by the UNSW Human Research Ethics Advisory Panel (File 3309), and all participants provided electronic informed consent to participate.

### COVID-19 in Australia

During the week that the study was conducted, the COVID-19 virus was already in Australia, but infections were limited and were predominantly cases where individuals had contracted the virus outside of Australia (Australian Government Department of Health, 2020a; Worldometer, 2020). At the end of day 1 (March 2), there were 33 confirmed cases; the first death in Australia from the virus occurred on this day, as did the first reported community transmissions. This number had risen to 93 by March 9, with 3 deaths and 18 cases that were likely to be community transmissions (no history of recent travel). Ten of these cases were associated with the Diamond Princess repatriation flight from Japan, and are counted in the patients’ states of residence.

### Participants

In total, 3,086 people viewed the participant information page and consent form. Of these, 854 either did not consent or completed only some of the survey questions before discontinuing, and 2,232 submitted the survey. Nine responses were excluded because participants reported that they did not live in Australia, and 49 responses were incomplete (48 missing demographic information, 1 with less than half of all responses completed). This resulted in a final sample of 2,174 participants.

### Measures

#### Information

First, participants were asked how closely they had been following news about the outbreak (11-point scale from *not at all* to *very closely*), and sources of information about the outbreak (select all that apply: news media, social media, official government websites, family members, friends or colleagues, none of the above, and other with text entry option). Participants were also asked about extent to which they believe that scientists and other medical and health experts understand COVID-19 (11-point scale from *don’t understand at all* to *understand very clearly*), to assess perceived scientific uncertainty.

#### Perceived risk and worry

Next participants were asked five questions relating to their perceived risk from, and worry about COVID-19. The first question assessed how concerned or worried respondents were feeling about the possibility of a widespread outbreak in Australia (i.e. the virus spreading from person-to-person more like a typical cold or flu virus; 5-point scale: *not at all concerned, a little concerned, moderately concerned, very concerned, extremely concerned*). Perceived likelihood of an outbreak, and perceived likelihood of the individual catching the virus if there was an outbreak, were assessed on a visual analogue scale (VAS) from 0 (*not at all likely*) to 100 (*extremely likely*). Perceived behavioural control, i.e. how much an individual could personally do to protect themselves from catching the virus, was assessed on a VAS from 0 (*couldn’t do anything*) to 100 (*could do a lot*). Finally, perceived severity was assessed by asking how severe respondents thought their symptoms would be if they did catch COVID-19; response options were: *no symptoms, mild symptoms, moderate symptoms, severe symptoms, severe symptoms requiring hospitalisation*, and *severe symptoms leading to death*.

#### Knowledge

To assess knowledge (and possible misinformation) about COVID-19, participants were asked to respond to a series of statements about the COVID-19 coronavirus, and whether (to the best of their knowledge) these statements were true, false, or they were unsure of the answer. These items and their correct answers (at the time of the study) can be seen in Table 2. On the day the study was launched (March 2, 2020), Australia experienced its first local transmission, and first death from the virus. The number of correctly answered items was summed to generate a virus knowledge subscale score.

**Table 1.** Demographic characteristics of the sample with number (percentage) of respondents.

| Demographic Variables | Total N (%) |
| --- | --- |
| **Gender** |  |
| Male | 503 (23.1) |
| Female | 1635 (75.2) |
| Non-binary, different identity, or prefer not to say | 36 (1.7) |
| **State** |  |
| New South Wales | 934 (43.0) |
| Victoria | 312 (14.4) |
| Queensland | 387 (17.8) |
| South Australia | 122 (5.6) |
| Western Australia | 261 (12.0) |
| Tasmania | 87 (4.0) |
| Australian Capital Territory | 52 (2.4) |
| Northern Territory | 19 (0.9) |
| **Age Group** |  |
| 18-29 | 489 (22.5) |
| 30-49 | 857 (39.4) |
| 50-59 | 487 (22.4) |
| 60+ | 303 (13.9) |
| Not stated | 38 (1.7) |
| **Ethnicity** |  |
| Caucasian (White / European) | 1639 (75.4) |
| Australian Aboriginal or Torres Strait Islander | 178 (8.2) |
| Asian | 173 (8.0) |
| Other or prefer not to say | 184 (8.5) |
| **Highest Education** |  |
| High school only: completed (Year 12) or not completed (Year 11 or below) | 534 (24.6) |
| Trade certificate, diploma, or advance diploma | 528 (24.3) |
| Bachelor's degree | 562 (25.9) |
| Graduate diploma, graduate certificate, or postgraduate degree | 543 (25.0) |
| Not stated | 7 (0.3) |

**Table 2.** Percentage of true, false, and unsure responses to general knowledge items, with correct answers in bold font

|  | True | False | Unsure |
| --- | --- | --- | --- |
| Currently there is no vaccine to protect against COVID-19 coronavirus [T] | **95.0** | 1.6 | 3.3 |
| There is an effective medicine available for treating COVID-19 coronavirus [F] | 5.1 | **79.2** | 15.6 |
| There are ways to help slow the spread of COVID-19 coronavirus [T] | **89.7** | 4.0 | 6.2 |
| If COVID-19 coronavirus breaks out in Australia, it is likely that some people will have natural immunity to it [F] | 29.5 | **34.1** | 36.3 |
| The ordinary flu vaccine will protect me from COVID-19 coronavirus [F] | 0.6 | **92.7** | 6.6 |
| To date, no one in Australia has died from COVID-19 coronavirus [F*] | 4.5 | **91.8** | 3.6 |
| To date, no one in Australia who was infected with COVID-19 coronavirus passed it on to infect another person [F*] | 4.1 | **84.9** | 10.9 |
| There are other strains of coronaviruses that can infect humans, including those that cause the common cold [T] | **80.2** | 4.7 | 15.0 |
| The health effects of COVID-19 coronavirus appear to be more severe for people who already have a serious medical condition [T] | **97.7** | 0.8 | 1.4 |
| Antibiotics are an effective treatment for COVID-19 coronavirus [F] | 3.4 | **81.9** | 14.5 |
| Packages or letters from China can spread the virus [F] | 6.8 | **67.8** | 25.3 |
| Taking vitamin C or other vitamins will protect you from the COVID-19 coronavirus [F] | 5.9 | **74.1** | 19.7 |
| There is no evidence that vaccines against pneumonia will protect you against the COVID-19 coronavirus [T] | **67.0** | 5.5 | 27.4 |
| Regularly rinsing your nose with saline will protect you against the COVID-19 coronavirus [F] | 2.7 | **77.6** | 19.6 |
| There is no evidence that eating garlic will protect you against the COVID-19 coronavirus [T] | **82.6** | 6.7 | 10.6 |
| Putting sesame oil on your body will block the COVID-19 coronavirus from entering your body [F] | 0.3 | **95.4** | 4.2 |
| Hand dryers are effective in killing the COVID-19 coronavirus [F] | 2.8 | **80.2** | 17.0 |
| The virus was genetically engineered as part of a biological weapons program [F] | 10.2 | **57.6** | 32.0 |
| The virus was human-made and deliberately released [F] | 10.2 | **57.8** | 31.9 |

\* True during study design, false at data collection
Note: missing data from 1-5 respondents for each item, percentages do not always total 100

To further assess the accuracy of their knowledge about the virus, participants were asked to identify the most common symptoms of COVID-19 infection, which was based on information provided to the Australian public at the time: fever, cough, sore throat, and shortness of breath (Australian Government Department of Health, 2020c). More recent versions of information provided includes fatigue or tiredness, which were not included in the survey. Three uncommon symptoms were included: diarrhea, vomiting, and nausea (Guan et al., 2020). The number of correctly answered items was summed to generate a symptoms knowledge subscale score. An additional knowledge assessment asked about the ways the virus can potentially be spread (see Table 3), including droplets spread through coughing or sneezing, touching or shaking hands with someone who is infected, and touching surfaces that have come into contact with the virus. Three other sources, which to date do not appear to be transmission mechanisms, were also included: water, mosquitos, and airborne spread (Centers for Disease Control and Prevention, 2020; World Health Organization, 2020b, 2020a). As above, the number of correctly answered items was summed to generate a transmission knowledge subscale score.

**Table 3.** Percentage of yes, no, and unsure responses to symptoms and transmission knowledge items, with correct answers in bold font

|  | Yes (%) | No (%) | Unsure (%) |
| --- | --- | --- | --- |
| **Symptoms** |  |  |  |
| Fever | **97.5** | 0.9 | 1.5 |
| Cough | **96.7** | 1.1 | 2.0 |
| Sore throat | **86.1** | 4.0 | 9.7 |
| Shortness of breath | **90.4** | 2.7 | 6.9 |
| Nausea | 20.9 | **50.4** | 28.4 |
| Vomiting | 8.5 | **64.4** | 26.8 |
| Diarrhoea | 13.9 | **66.0** | 19.7 |

| **Transmission** |  |  |  |
| --- | --- | --- | --- |
| Droplets spread through coughing or sneezing | **98.8** | 0.3 | 0.9 |
| Surfaces recently touched by someone who is sick | **91.2** | 3.1 | 5.7 |
| Touching or shaking hands with a person who is sick | **94.8** | 2.1 | 3.1 |
| Airborne | 56.1 | **28.1** | 15.8 |
| Waterborne | 8.0 | **64.4** | 27.3 |
| Mosquitos | 1.7 | **80.0** | 18.0 |

Finally, one item assessed knowledge of recommended face mask use (to minimise transmission of the virus, who should be wearing a face mask?), with the advice to the public being that people who were sick should be wearing masks to stop them spreading the virus, but that masks were not recommended for healthy people (Australian Government Department of Health, 2020c). A final item assessed knowledge of the approximate mortality rate, which at the time was a figure reported by WHO as 3.4% (World Health Organization, 2020d). Responses were deemed correct if they were between 1 and 5%. A total COVID-19 knowledge score was calculated as the number of correct responses to all items assessing various aspects of knowledge about COVID-19, potentially ranging from 0 to 34.

### Health-protective behaviours

To assess distancing and hygiene behaviours, participants were asked whether they had engaged in a total of 14 behaviours during the previous month, and whether they would engage in any of these behaviours in the case of a more widespread outbreak in the future (see Table 4). Response options for both questions were *yes, no, unsure*, and *not applicable*. Items were generated based on previous research (Bults, Beaujean, Richardus, & Voeten, 2015; Petrie et al., 2016; Rubin et al., 2009; Simpson et al., 2019) and recommended behaviours (Australian Government Department of Health, 2020c). One additional item (‘reduce or avoid going to Chinese restaurants, Chinatowns, or Chinese precincts’) was included to assess the prevalence of this reaction following media reports indicating public avoidance (Johanson, Gross, & Cavataro, 2020). Health-protective behaviour sum scores (number of ‘yes’ responses) were calculated separately for current behaviours and intended future behaviours should an outbreak occur, excluding the item asking about Chinese restaurants or precincts, thus possible scores range from 0 to 13.

**Table 4.**
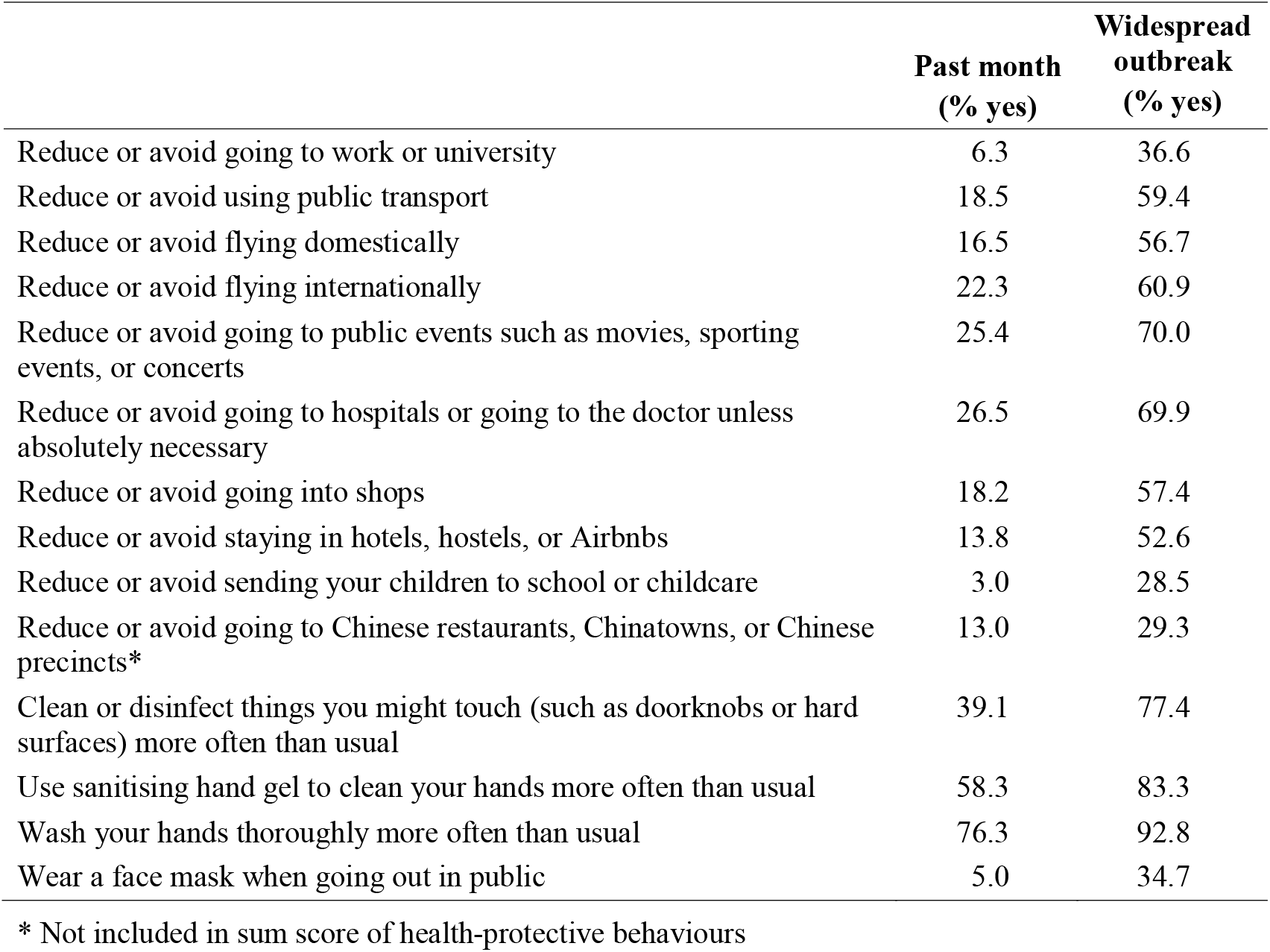
Percentage of yes responses relating to health-protective behaviours during the past month, and anticipated behaviours during a widespread outbreak

Finally, participants were asked to complete a single item asking about how likely it is that they would choose to have a vaccination for the COVID-19 coronavirus, if there was a safe and effective vaccine developed. Response options were on a 5-point scale: *would definitely not get the vaccine, would probably not get the vaccine, unsure if I would get the vaccine or not, would probably get the vaccine, would definitely get the vaccine*. Responses were scored such that higher scores indicated higher vaccine intentions.

#### Demographics and health information

Information was collected on participants’ age group, gender, ethnicity, highest level of education, and region of residence within Australia. Demographic characteristics of the sample can be seen in Table 2. In addition, participants were asked to complete three questions relating to their health. First, a single-item measure assessing their self-rated heath (Idler & Benyamini, 1997), with responses on a 5-point scale from *poor* to *excellent*. Second, an item assessing whether that had received a flu vaccine in the previous year (*yes, no, unsure*). For the purposes of analysis, *no* and *unsure* responses were combined to form a dichotomous measure. Finally, participants were asked whether they, or any family members or friends, had caught COVID-19 (*yes, no, unsure*). Only 9 respondents said ‘yes’ to this question, and these responses were included in the analysis.

## Results

### Demographics

Demographic characteristics of the sample can be seen in Table 1. A large proportion of respondents were from the state of New South Wales (NSW). In part this is likely because of the association of the study with the University of New South Wales. It may also reflect that, at the time of the survey, the majority of COVID-19 cases (51% on the final day of data collection) were in NSW and thus people in the state had heightened awareness and greater willingness to participate in the research.

#### Health-related characteristics

Respondents’ self-rated health was measured on a scale from poor (1) to excellent (5), with a mean of 3.21 (*SD* = 0.98). The majority of participants rated their health as good (38.7%) or very good (29.5%), and a smaller proportion rated their health as excellent (9%). Relatively few respondents rated their health as poor (4.3%), and a 18.2% rated their health as fair. Approximately half of the sample (52.9%) reported having had a flu vaccine in the past year, while 45.8% had not had a flu vaccine, 1.1% were unsure, and 0.2% (4 respondents) declined to answer. Only 9 respondents (0.4%) reported that they themselves, or their friends or family, had caught COVID-19. The majority responded had not (95.3%), and 90 (4.1%) were unsure.

### Information

Participants were asked how closely they had been following news about COVID-19 on a scale from 0 (not closely at all) to 10 (very closely). The majority (68.4%) rated their news engagement as 7 or higher, and 18.5% rated their engagement as a 10. The mean score on this scale was 7.3 (*SD* = 2.1). Participants reported getting information about COVID-19 from the news media (85.2%), official government websites (72.2%), social media (68.5%), colleagues or friends (22.7%), and family members (22.7%). ‘Other’ sources of information were indicated by 15.6% (n = 340) of respondents, with the most common open-ended responses being workplace communication (n = 57; 2.6%), medical and academic journals or publications (n = 53; 2.4%), Reddit (n = 45; 2.1%), directly from health professionals (n = 29; 1.3%), and YouTube videos (n = 28; 1.3%). Only 0.3% respondents reported not getting information from any of these sources.

Perceived scientific uncertainty – the extent to which respondents report believing that scientists and other medical and health experts understand COVID-19 – was moderate. The mean rating on a scale from 0 (don’t understand at all) to 10 (understand very clearly) was 6.1 (*SD* = 2.0).

### Perceived Risk

Concern about the possibility of a widespread outbreak in Australia was typically moderate (*M* = 3.2, *SD* = 1.1; scale from 1 to 5, where 1 = not at all, 5 = extremely concerned). A small proportion reported being not at all concerned (6.1%), while 24% reported being a little concerned, 31.1% were moderately concerned, 21.7% were very concerned, and 14.9% extremely concerned. Respondents’ ratings of the perceived likelihood of an outbreak of COVID-19 in Australia was relatively high (*M* = 71.8, *SD* = 24.9; scale from 0 to 100), and perceived likelihood that they would catch the virus in the case of an outbreak was moderate (*M* = 54.9, *SD* = 24.7). Perceived behavioural control, or the belief that personal protective behaviours could help prevent infection, had a mean score of 68.2 (*SD* = 21.6).

With regard to perceived severity of symptoms in the case of infection, only 0.3% of respondents indicated that they would experience no symptoms; mild (27.5%) and moderate (46.7%) symptoms were most commonly anticipated. However, one in four respondents perceived the illness severity to be high: with 14.1% indicating they thought they would experience severe symptoms, severe symptoms requiring hospitalisation (8.8%), or severe symptoms leading to death (2.3%).

### Knowledge

Participants were asked to respond to a series of true-false questions to assess their more general knowledge of COVID-19. The percentage of true, false, and unsure responses (with correct answers in bold font) can be seen in Table 2. Total general knowledge subscale scores were calculated out of 19 total items. Scores ranged from 0 (1 respondent) to 19 (129 respondents), with a mean of 14.9 (*SD* = 2.8).

Knowledge questions were also asked relating to most common symptoms and routes of transmission (see Table 3), with correct answers also summed to generate symptoms and transmission subscale scores. Respondents were more accurate in recognising the symptoms that have been linked with COVID-19, and less certain of whether the other symptoms (nausea, vomiting, diarrhoea) were indicative of illness. Symptoms knowledge subscale scores ranged from 0 to 7, with 32.6% of respondents correctly answering every item. The mean subscale score was 5.5 (*SD* = 1.4) indicating good recognition of the symptoms commonly mentioned in public health information provided to the Australian public at this time. Similarly, respondents typically recognised transmission routes associated with droplet spread, but were less certain of whether the virus can also spread via air, water, or mosquitos (current evidence indicates that it cannot). Transmission knowledge subscale scores ranged from 0 to 6, with a mean of 4.6 (*SD* = 1.0). Only 17.8% of respondents correctly answered every item.

An additional item assessed knowledge of recommended face mask use. Most respondents (79.7%) correctly identified that people who were sick should wear masks to stop them spreading the virus. However, 15.9% incorrectly reported that ‘everyone’ – both sick and healthy – should be wearing masks, and 1.3% responded that only healthy people should be wearing masks. In a few cases (2.7%) respondents said that ‘no one’ should be wearing a face mask, and 7 people (0.3%) did not respond to this question. A final item assessed knowledge of the approximate mortality rate, and 69.2% of respondents gave answers between 1 and 5%, which were deemed accurate. Percentage estimates ranged from 0 (0.5%) to 100 (0.3%), with a mean of 7.84% (*SD* = 12.31). A total COVID-19 knowledge score was calculated from responses to general, symptoms, and transmission subscales, and individual items about mask use, and mortality. Scores ranged from 7 to 34 (out of a possible 34), with a mean of 26.48 (*SD* = 4.10).

### Health-Protective Behaviours

The percentage of respondents who reported having engaged in a range of distancing and hygiene behaviours during the past month, and the percentage who reported intending to engage in these behaviours in the case of a widespread outbreak in Australia, can be seen in Table 4. During the previous month, hygiene behaviours (hand washing, using hand sanitising gel, and cleaning and disinfecting surfaces) were the most commonly reported behaviours. Respondents also intended to engage in these hygiene behaviours in a widespread outbreak, as well as distancing behaviours including avoidance of public events, domestic and international travel, and using public transport.

The number of behaviours endorsed for each timeframe was summed. Scores for behaviours in the past month ranged from 0 (16%) to 13 (0.3%), with most (80.5%) of respondents reporting five behaviours or fewer, and a mean score of 3.29 (*SD* = 2.89). Respondents anticipated engaging in more distancing and hygiene behaviours during a widespread outbreak (M = 7.78, SD = 3.65), with only 2.5% reporting that they would not engage in any of the listed behaviours, and over one third (38.7%) indicating that they would engage in 10 or more behaviours.

Finally, participants were asked whether they would choose to have a vaccination for the virus if a safe and effective vaccine was developed. Four in five respondents indicated that they would either definitely (60.4%) or probably (20.8%) get the vaccine. Only 12.3% reported being unsure, 3.7% said that they would probably not get the vaccine, and 2.8% said that they would definitely not get vaccinated against the COVID-19 coronavirus.

### Predictors of Health-Protective Behaviours

Negative binomial regressions with maximum likelihood estimation were conducted to assess the influence of information, perceived risk, and knowledge-related predictors on engagement (past month and anticipated in the case of a widespread outbreak) with health-protective behaviours, while controlling for demographic factors and self-rated health. Negative binomial regression was chosen for health-protective behaviours because it is appropriate for over-dispersed count data, and the health-behaviour outcome measures are both counts of the number of behaviours endorsed, and are over-dispersed as the variance of measures exceeds the mean score.

#### Demographic predictors

First, to assess demographic differences in health-protective behaviours, each demographic predictor variable was entered individually into a separate negative binomial regression model. The mean (standard error) number of behaviours engaged in or anticipated across demographics can be seen in Table 5. For behaviours during the past month, demographic differences were seen by gender (*p* < .001), state of residence (*p* = .002), age group (*p* = .001), and ethnicity (*p* < .001). Female respondents reported engaging in more health-protective behaviours than their male counterparts, and those in the youngest age group (18-29) engaged in fewer behaviours than older respondents. Behaviour differences by ethnicity were also seen, with non-Caucasian respondents reporting more health-protective behaviours. Respondents from Queensland reported engaging in more behaviours than those from the category of New South Wales (reference category). There was not a significant effect of education level (*p* = .339).

**Table 5.** Demographic differences in the mean (SE) number of health-protective behaviours over the past month, and anticipated behaviours in the case of a widespread outbreak.

| Demographic Variables | Past month<br>M (SE) | Widespread<br>outbreak<br>M (SE) |
| --- | --- | --- |
| **Gender** |  |  |
| Male (RC) | 2.83 (0.12) | 7.56 (0.17) |
| Female | 3.45 (0.08)* | 7.86 (0.10) |
| Non-binary, different identity, or prefer not to say | 2.75 (0.43) | 7.00 (0.61) |
| **State** |  |  |
| New South Wales (RC) | 3.15 (0.09) | 7.79 (0.13) |
| Victoria | 3.08 (0.16) | 7.35 (0.21) |
| Queensland | 3.86 (0.17)* | 8.31 (0.21)* |
| South Australia | 2.70 (0.23) | 7.01 (0.33)* |
| Western Australia | 3.40 (0.19) | 7.79 (0.24) |
| Tasmania | 3.47 (0.33) | 7.74 (0.42) |
| Australian Capital Territory | 3.38 (0.42) | 7.85 (0.42) |
| Northern Territory | 3.58 (0.73) | 8.47 (0.96) |
| **Age Group** |  |  |
| 18-29 (RC) | 2.86 (0.12) | 7.47 (0.17) |
| 30-49 | 3.52 (0.11)* | 8.35 (0.14)* |
| 50-59 | 3.25 (0.13)* | 7.50 (0.17) |
| 60+ | 3.32 (0.17)* | 7.15 (0.21) |
| **Ethnicity** |  |  |
| Caucasian (White / European; RC) | 3.03 (0.07) | 7.51 (0.09) |
| Australian Aboriginal or Torres Strait Islander | 3.85 (0.25)* | 8.44 (0.31)* |
| Asian | 4.74 (0.30)* | 9.17 (0.34)* |
| Other or prefer not to say | 3.69 (0.24)* | 8.18 (0.30)* |
| **Highest Education** |  |  |
| High school only: completed (Year 12) or not completed (Year 11 or below; RC) | 3.43 (0.13) | 7.66 (0.17) |
| Trade certificate, diploma, or advance diploma | 3.39 (0.13) | 7.92 (0.17) |
| Bachelor's degree | 3.16 (0.12) | 7.80 (0.17) |
| Graduate diploma, graduate certificate, or postgraduate degree | 3.19 (0.12) | 7.72 (0.17) |

RC: Reference Category
\*Significantly different from the Reference Category at .05

For anticipated behaviours during a widespread outbreak, demographic differences were seen by state (*p* = .023), age (*p* < .001), and ethnicity (*p* < .001). Respondents from Queensland reported intending to engage in more behaviours, and those from South Australia intending to engage in fewer than those in New South Wales. Those in the 30-49 age bracket reported intending to engage in more health-protective behaviours during an outbreak compared to younger respondents. Similar to the results from behaviours over the previous month, intended behaviours during an outbreak were higher in groups of non-Caucasian respondents. There was no longer a significant effect of gender (*p* = .153), and education remained non-significant (*p* = .738).

#### Psychological predictors of health-protective behaviours during the past month

To assess the influence of psychological predictors on engagement with health-protective behaviours during the past month, all relevant variables were entered into a single model (see Table 6), controlling for demographic variables and self-rated health. The Pearson Chi-Square Goodness of Fit statistic (1.084) indicated that the model fit the data well. The omnibus test results indicate that the model was a significant improvement over a null model, *χ*^2^ = 940.41 (df = 2) *p* < .001.

**Table 6.**
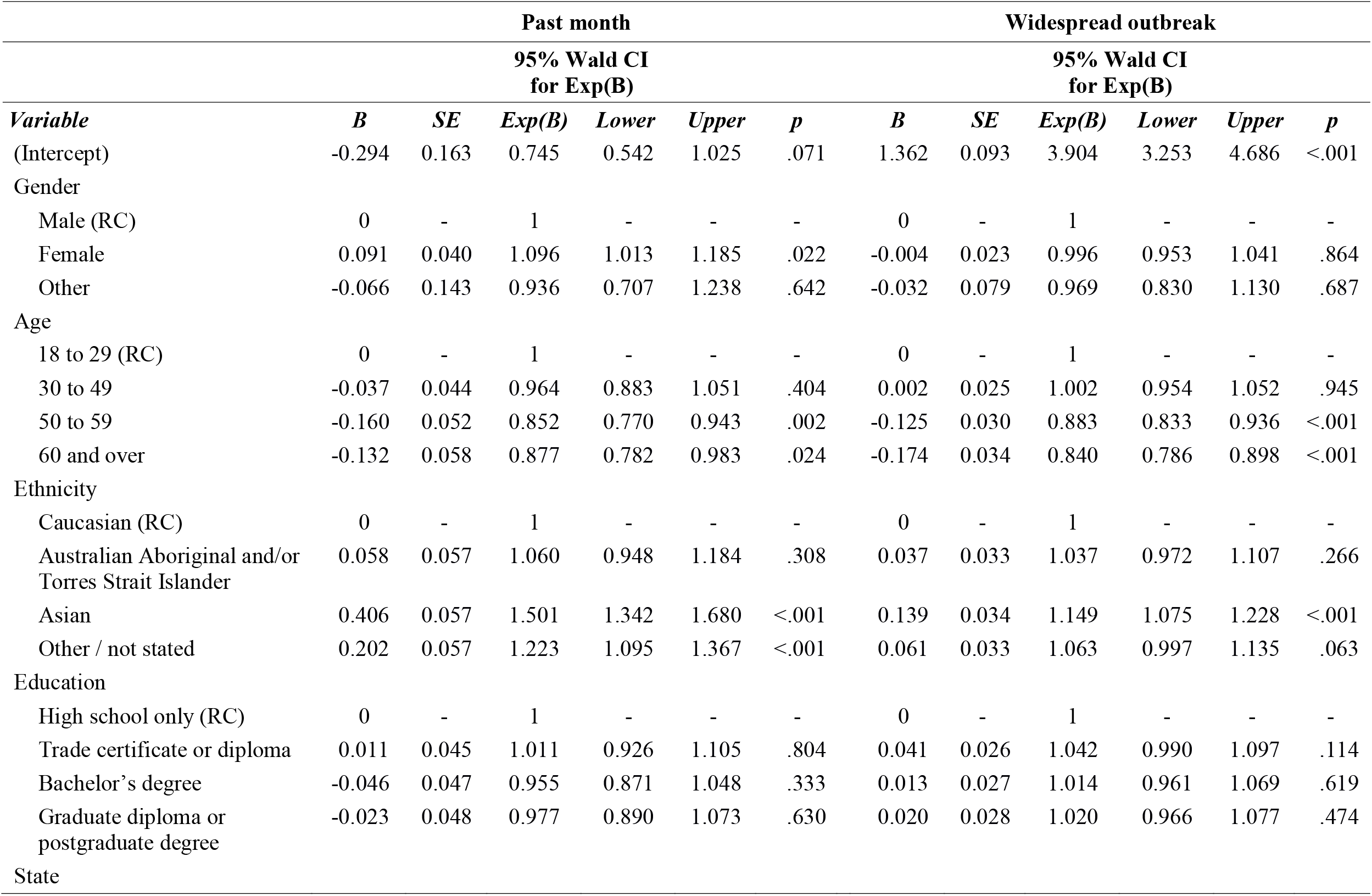
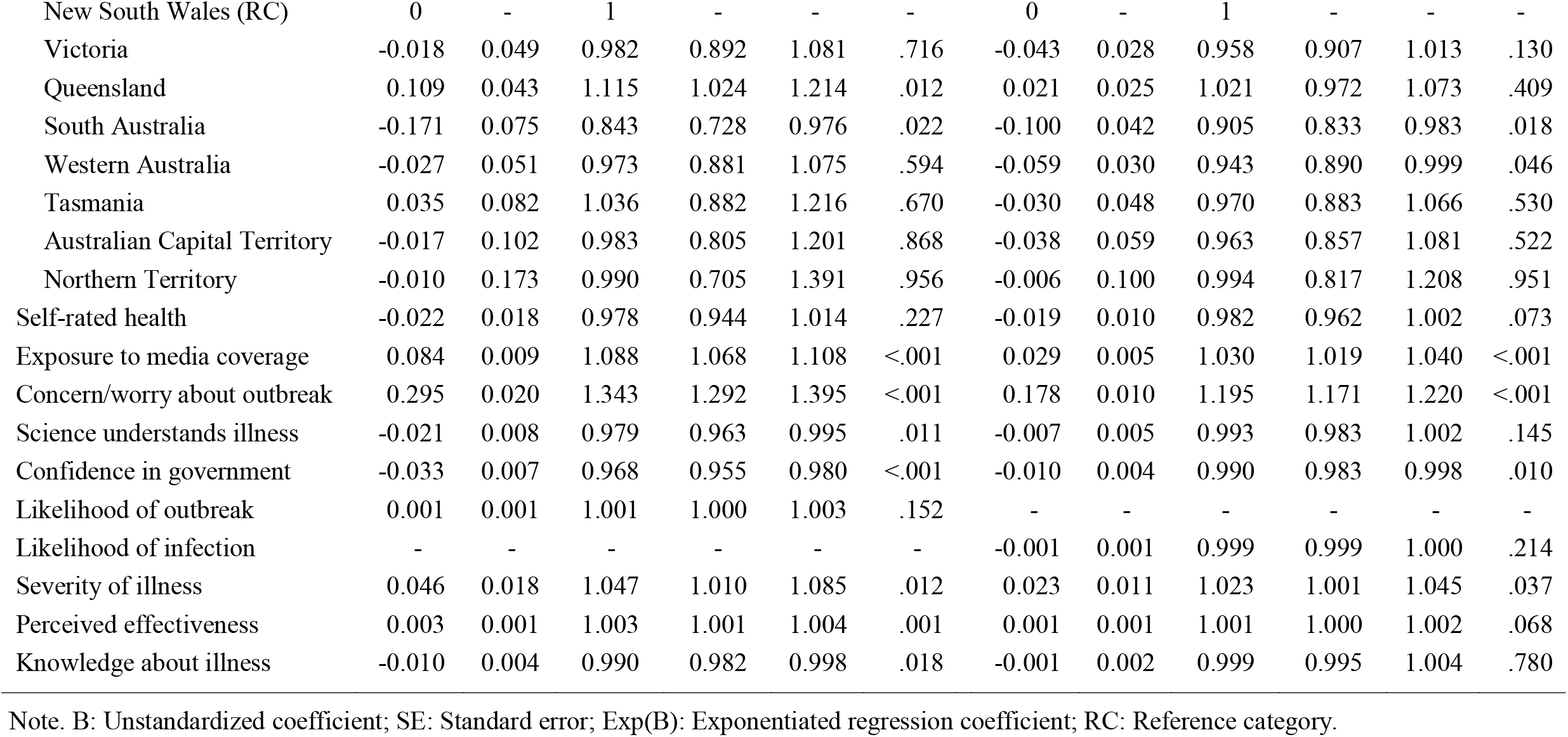
Predictors of the number of health-protective behaviours during the past month (left), and anticipated health-protective behaviours during a widespread outbreak (right).

More closely following media coverage, heightened worry or concern about an outbreak, higher perceived personal severity of COVID-19, and higher perceived effectiveness of health-protective behaviours were significant predictors of heightened engagement with distancing and hygiene behaviours during the previous month. In contrast, stronger beliefs in scientific and medical understanding of the virus, confidence in government information, and higher COVID-19 knowledge scores, predicted reduced engagement with health-protective behaviours during the month prior to completing the survey.

#### Anticipated health protective behaviours during a widespread outbreak

The analysis for anticipated future outbreak behaviours was almost identical to that for past month behaviours – except that the item assessing perceived likelihood of an outbreak was removed, and likelihood of infection in the case of an outbreak was added in its place. All variables were entered into a single model (see Table 6). The Pearson Chi-Square Goodness of Fit statistic (1.083) indicated good model fit. The omnibus test results indicated that the model was a significant improvement over a null model, χ^2^ = 698.73 (df = 27) *p* < .001.

Similar to the psychological predictors of current behaviours, the number of anticipated behaviours during a widespread outbreak was predicted by more closely following media coverage, higher concern or worry about an outbreak, and perceived seriousness of COVID-19 symptoms. However, perceived effectiveness of protective behaviours was no longer a significant predictor. Similar to the results for current behaviours, greater perceptions of scientific and medical understanding of the virus predicted reduced intentions to engage in health-protective behaviours in the case of a widespread outbreak. COVID-19 knowledge scores were not associated with intended behavioural responding.

### Predictors of Vaccination Intentions

Finally, respondents were asked how likely they were to get vaccinated against COVID-19 if a safe and effective vaccine was developed (see Table 7 for response options). Because this outcome was not a count variable, and did not approximate a normal distribution, ordinal logistic regression was used to assess the influence of information, perceived risk, and knowledge-related predictors on vaccination intentions, while controlling for demographic factors and self-rated health.

**Table 7.** Number and percent of respondents in each age group reporting that they would definitely not, probably not, were unsure, probably would, or definitely would get a COVID-19 vaccine, if available.

|  | N (% of age group) |  |  |  |  |
| --- | --- | --- | --- | --- | --- |
|  | 18-29 | 30-49 | 50-59 | 60+ | Total |
| Definitely not | 6 (1.2) | 24 (2.8) | 17 (3.5) | 13 (4.3) | 60 (2.8) |
| Probably not | 14 (2.9) | 31 (3.6) | 22 (4.5) | 11 (3.6) | 78 (3.7) |
| Unsure | 44 (9.0) | 118 (13.8) | 76 (15.6) | 28 (9.2) | 266 (12.5) |
| Probably would | 133 (27.2) | 190 (22.2) | 74 (15.2) | 42 (13.9) | 439 (20.6) |
| Definitely would | 292 (59.7) | 494 (57.6) | 297 (61.1) | 209 (69.0) | 1292 (60.5) |

#### Demographic predictors

First, to assess demographic differences in vaccine intentions, each demographic predictor variable was entered individually into a separate ordinal logistic regression model. There were no demographic differences in vaccine intentions by gender (*p* = .429), state of residence (*p* = .832), ethnicity (*p* = .461), or level of education (*p* = .129). Respondents did differ in their vaccine intentions by age group (*p* = .019). Compared to those in the 60 plus age group, being in the 30-49 (*ExpB* = 0.662, 95%CI [0.503 to 0.871], *p* = .003) or 50-59 (*ExpB* = 0.695, 95%CI [0.515 to 0.938], *p* = .017) age group was associated with a lower likelihood of intending to get a vaccination if one becomes available (see Table 7 for percent of responses in each category by age group).

#### Psychological predictors of vaccination intentions

Predictors entered into the full model were the same as in the previous analysis, with the addition of a dichotomous variable reflecting whether respondents had received a flu vaccine in the previous year, or had not / were unsure. All variables were entered into a single model (see Table 8). The Pearson Chi-Square Goodness of Fit statistic (0.921) indicated good model fit. The omnibus test results indicate that the model is a significant improvement over a null model, *χ*^2^ = 557.23 (df = 28) *p* < .001.

**Table 8.**
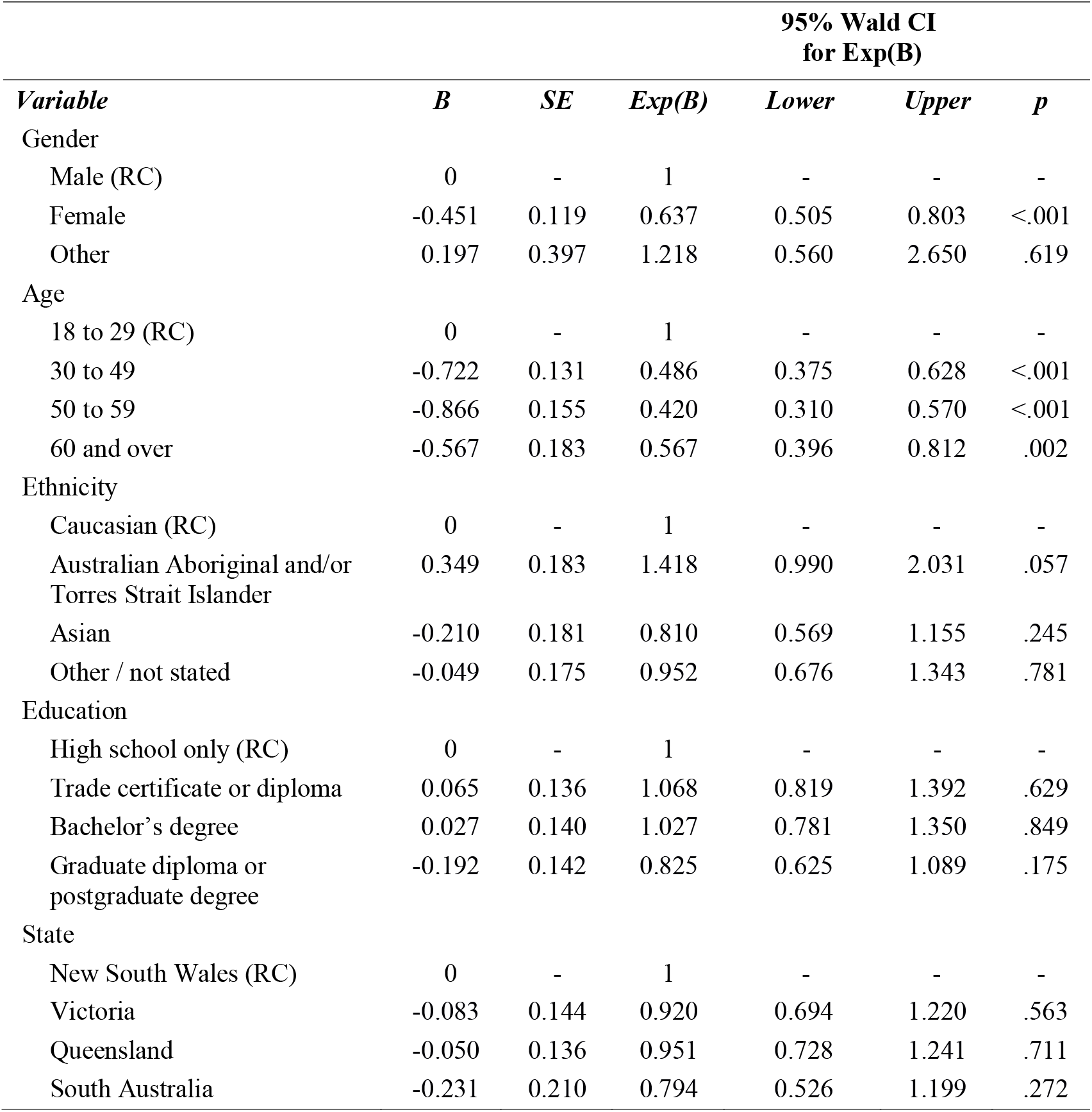
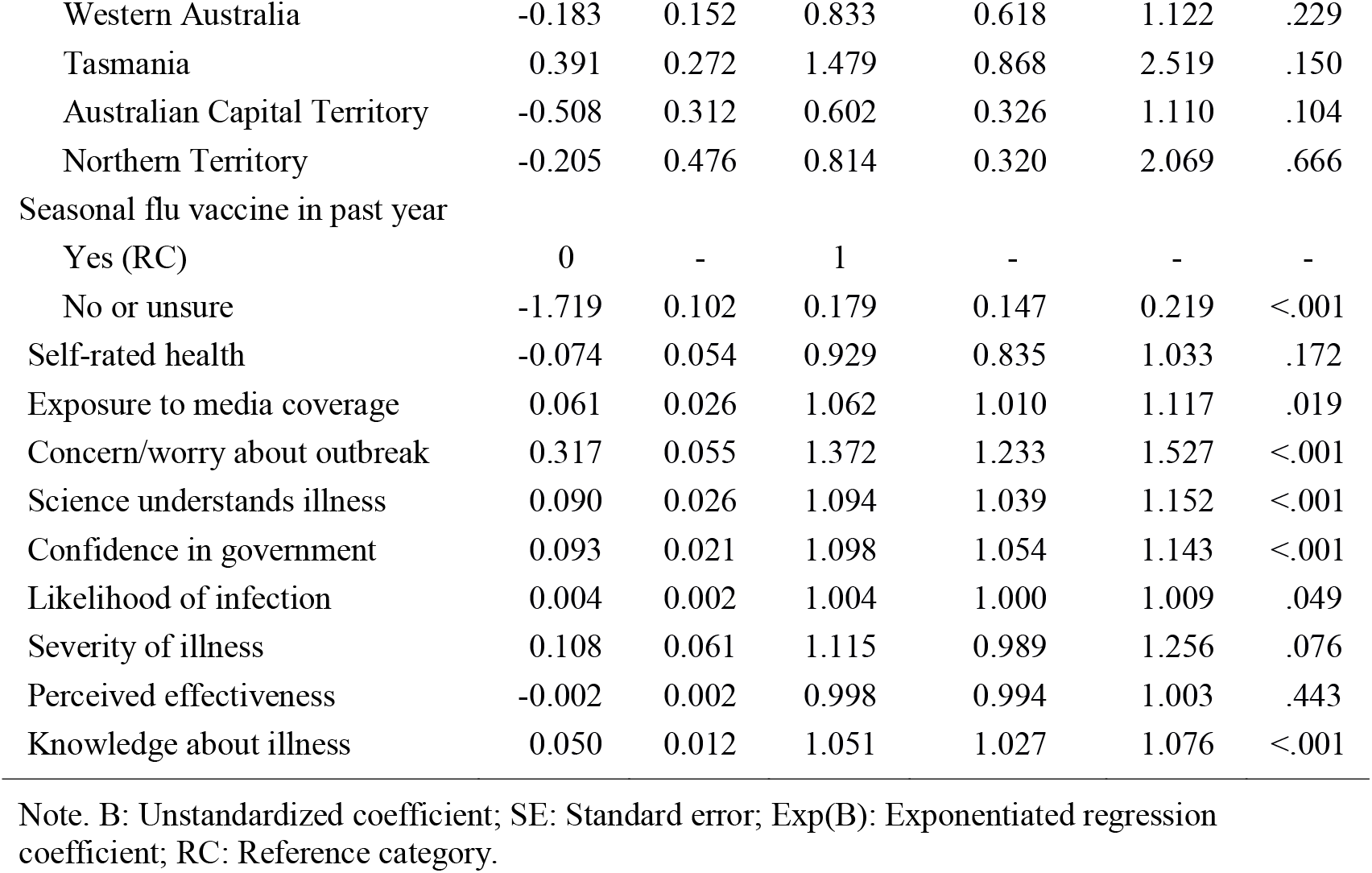
Predictors of likelihood of getting vaccinated against COVID-19 if a vaccine becomes available.

Having received a seasonal flu vaccine in the past year predicted increased intentions to get a COVID-19 vaccine if it becomes available. With regard to psychological predictors and in line with previous results, both increased exposure to media coverage and heightened worry or concern about the outbreak predicted increased vaccination intentions. In contrast to results relating to health-protective behaviours, perceptions of greater scientific and medical understanding of the virus, confidence in government information, and higher knowledge scores, predicted greater vaccination intentions.

## Discussion

The results of the survey provide the first information on public knowledge, perceived risk and worry, and health-protective behaviours in the early period of the COVID-19 pandemic in Australia, in a sample of 2174 participants recruited during the first week of March 2020 (2-9^th^), shortly before more widespread community transmission. Overall, just over one third of participants were very or extremely worried or concerned about the possibility of a widespread outbreak in Australia, and a further third reported moderate worry. In line with this finding, the perceived likelihood of an outbreak was relatively high, with 74% of participants giving a rating of 60 or higher (on a scale from 0 to 100, where 100 was extremely likely), and 19% giving a rating of 100. Perceived personal risk of catching the virus in the case of an outbreak was more moderate, with 44% of respondents rating this as 60 or higher. Consistent with the findings from past pandemics such as SARS (Bults et al., 2011), higher worry about outbreaks was associated with greater actual health-protective behaviours (e.g., handwashing), as well as anticipated behaviours in the event of a future outbreak. Recent research from China regarding the COVID-19 outbreak indicates that engaging in hand hygiene and other health-protective behaviours was associated with reduced psychological impact of the outbreak, including lower stress and anxiety (Wang et al., 2020). These findings highlight the importance of encouraging the public to engage with such behaviours not only to reduce the risk of infection, but also to reduce the psychological impact of the pandemic.

This study provided important new knowledge about what participants *expected* in terms of how serious the symptoms of coronavirus would be, should they contract COVID-19. There is a clear discrepancy between respondents’ perceived severity of symptoms and current data on rates of asymptomatic infection. Only 0.3% of respondents believed that they would experience no symptoms, whereas mild (28%) and moderate symptoms (47%) were expected. In contrast, emerging evidence from groups with widespread testing for the SARS-CoV-2 virus (e.g. cruise ships, repatriation flights, and overseas arrivals) indicates that between 2 and 8 out of every 10 infections have been asymptomatic (Day, 2020; Mizumoto, Kagaya, Zarebski, & Chowell, 2020; Nishiura et al., 2020). In addition, the incubation period for the SARS-CoV-2 virus is approximately five days (Lauer et al., 2020). Although those in the incubation period and with asymptomatic infections experience no symptoms of illness, they are still able to transmit the virus to others (Bai et al., 2020; Zou et al., 2020). People commonly rely on symptoms to indicate illness, and assume that the absence of symptoms means they are well (Diefenbach & Leventhal, 1996). Such assumptions in the COVID-19 pandemic could have serious consequences, both in terms of community transmission, and reduced health-protective behaviours both by those with asymptomatic infections and those close to them. Therefore, public health communication campaigns about COVID-19 need to address these misconceptions.

The results also provide important information about where Australian residents are seeking their information about COVID-19, and how accurate their knowledge is about the virus and its transmission. While it was promising to see that 72% sourced information from official and government websites, mainstream news media was most popular (85%), and social media (68%) was also high. The high usage of news media as sources of information is concerning given the potential for alarming, sensationalist portrayals of the pandemic (Klemm et al., 2016). In addition, myths, rumours and misinformation can quickly spread online, particularly via social media. People are more likely to spread false news stories and misinformation, and these false stories spread more widely, and more quickly, than true stories (Vosoughi, Roy, & Aral, 2018). This might have been one of the reasons why there were several uncertainties around COVID-19, with a significant minority reporting being ‘unsure’ whether people have natural immunity to it, whether specific home remedies (garlic, vitamins, rinsing noses with saline) help protect against coronavirus. It may also explain some uncertainty around whether packages and letters from China spread the virus, and whether the virus was human made and deliberately released; almost one third of respondents indicated feeling ‘unsure’ whether this was true or not. In addition, uncertainty and rapidly changing information may have contributed to increased worry about the virus (Han, Moser, & Klein, 2006). These findings speak to the importance of distributing accurate health information about COVID-19 through a variety of sources (news, social media, government websites) to reach the general population and correct misinformation.

Only one third of participants correctly answered all of the questions about the symptoms of COVID-19, and less than 20% correctly answered all questions around transmission, suggesting more education of the general public about COVID-19 is needed. Overall, respondents had good knowledge of the symptoms most commonly associated with infection, and understood that transmission appears to be primarily through respiratory droplets. Over 95% of respondents correctly identified fever and cough as COVID-19 symptoms, and over 90% correctly identified droplet spread (including droplets on hands or surfaces) as the primary route of transmission. There were a few uncertainties around whether gastrointestinal symptoms were typically associated with COVID-19, though evidence indicates these symptoms are relatively are uncommon (Guan et al., 2020). Similarly, there was uncertainty around airborne, waterborne, and mosquito transmission. Waterborne and mosquito transmission are highly unlikely (Centers for Disease Control and Prevention, 2020; World Health Organization, 2020a). Although airborne transmission may be possible in some specific medical procedures (e.g. disconnecting a patient from a ventilator), available evidence does not indicate that airborne community transmission is common (World Health Organization, 2020b). However, uncertainties about routes of transmission, particularly uncertainty over whether the virus may be airborne, is likely to be concerning for members of the public.

The way people act during a pandemic is critical in lowering the risk of contracting and spreading the virus. Given the rapidly evolving situation with COVID-19 globally, the findings from this study (which were assessed during the first week of March 2020) may not be reflective of actual behaviours now that higher restrictions have been put in place, and significant widespread messaging around social distancing, handwashing and self-isolation has been disseminated. However, our findings provide important insights into the demographic and psychological predictors of actual health-protective behaviours and anticipated health protective behaviours in the early stages of a pandemic disease outbreak. The most powerful predictors were demographic factors, including younger age, female gender, and being of non-Caucasian ethnicity, as well as risk perceptions (greater concern about outbreak and perceived severity of illness), and higher media exposure. The effect of media exposure is likely to be related to the provision of important health information about the pandemic. Although media exposure early in the outbreak appears to have facilitated health-protective behaviours, media fatigue – where people become desensitised to ongoing messaging – may reduce this effect as the pandemic continues (Collinson, Khan, & Heffernan, 2015). Repeated media exposure may also lead to heightened stress and anxiety which can have longer-term health effects, as well as contributing to excessive or misplaced health-protective behaviours such as presenting for diagnostic testing when actual risk of exposure is low (Garfin, Silver, & Holman, 2020).

The results of this study shed light on how many participants planned to get the COVID-19 vaccine if and when one becomes available; this information will be important for planning public health initiatives if and when a vaccine becomes available. A little over half of the sample (53%) had a flu vaccine in the past year, and prior flu vaccine predicted greater intentions to get a COVID-19 vaccine. Similar variables, including concern about outbreaks, greater media exposure, higher knowledge, and perceived scientific understanding predicted planned vaccination intentions. These findings are in line with previous research showing that concern about outbreaks and higher knowledge were associated with increased Ebola vaccine intentions (Petrie et al., 2016). In contrast with other research, perceived likelihood and severity of infection were only marginally associated with intentions to get a vaccine if one becomes available (Bish & Michie, 2010; Weinstein et al., 2007). In part this may be because the literature tends to focus on personal risk, yet in the case of COVID-19 the personal risk to most respondents is low, and behaviour may be driven primarily by concern for others. It may also be that because concern and worry was particularly high during data collection early in the outbreak (Bults et al., 2011), this affective response to the risk posed by the novel coronavirus was a particularly strong driver of both health-protective behaviours and vaccine intentions (Slovic et al., 2007).

The current study has limitations that should be taken into account. Participants were recruited online, primarily through Facebook, and therefore may not be representative of the general population. Although we recruited a range of education levels, cultural backgrounds, age ranges, and gender, three quarters were female, and Caucasian, 43% were from NSW, and 62% were younger than 50 years of age, and results may not generalise to the broader population. To maximise convenience sampling, we used solely self-report measures, which may lead to biased effects. While the results of the regression analyses provide interesting starting points to identify the demographic and risk variables that predict actual and anticipated health behaviours, as well as vaccine intentions, they cannot establish causality, and must be interpreted with caution. Given the large sample, the relationships between some of the significant predictors are likely to be small and may not be clinically meaningful.

The current results provide timely and important information on the Australian public responses to the COVID-19 pandemic, including information sources and engagement, knowledge, and perceived risk in the early stages of the outbreak in Australia, and their relationship with current and intended health-protective behaviours and vaccine intentions. Overall, the findings show that perceived risk of outbreak and personal risk of contracting COVID-19 were moderate immediately prior to a widespread outbreak, but there was a critical mismatch between expected severity of symptoms versus data on how COVID-19 is experienced, which need to be addressed in government education campaigns. Although health-protective behaviour was relatively low at the start of the outbreak, intended behaviours in the case of an outbreak were reassuringly high. These behaviours, and vaccination intentions were consistently predicted by greater exposure to media and worry about outbreaks. Finally, our questions revealed significant uncertainty and misinformation which needs to be corrected.

Without a vaccine currently available, encouraging widespread and sustained engagement with hygiene and distancing behaviours is critical to successfully managing the COVID-19 pandemic, flatten the curve of infections, and protect vulnerable individuals and overburdened healthcare systems. The results of the current study provide important insights into psychological and behavioural responses early in the outbreak of this novel coronavirus, and point to types of information and particular groups that may benefit from clear and targeted messaging to address misinformation and promote engagement with these health-protective behaviours.

## Data Availability

Ethical consent to make the data associated with this study available was not sought from participants. In addition, due to the low number of COVID-19 cases in Australia at the time data collection took place, making this data publicly available would potentially compromise the anonymity of those participants who reported that they themselves, or their friends or family, had been diagnosed with COVID-19.

## Author Contributions

KF and JN were responsible for the concept and design of the study, interpretation of results, writing and critical review of the manuscript. KF was responsible for data collection and analysis.

## References

Asmundson, G. J. G., & Taylor, S. (2020). Coronaphobia: Fear and the 2019-nCoV outbreak. Journal of Anxiety Disorders, 70(February). https://doi.org/10.1016/j.janxdis.2020.102196

Australian Government Department of Health. (2020a). Coronavirus (COVID-19) current situation and case numbers. Retrieved March 9, 2020, from https://www.health.gov.au/news/health-alerts/novel-coronavirus-2019-ncov-health-alert/coronavirus-covid-19-current-situation-and-case-numbers

Australian Government Department of Health. (2020b). Coronavirus (COVID-19) health alert. Retrieved March 7, 2020, from https://www.health.gov.au/news/health-alerts/novel-coronavirus-2019-ncov-health-alert#what-we-are-doing

Australian Government Department of Health. (2020c). What You need to know about coronavirus (COVID-19). Retrieved February 19, 2020, from https://www.health.gov.au/news/health-alerts/novel-coronavirus-2019-ncov-health-alert/what-you-need-to-know-about-coronavirus-covid-19

Bai, Y., Yao, L., Wei, T., Tian, F., Jin, D., Chen, L., & Wang, M. (2020). Presumed Asymptomatic Carrier Transmission of COVID-19. JAMA, 19–20. https://doi.org/10.1056/nejmoa2001316

Bish, A., & Michie, S. (2010). Demographic and attitudinal determinants of protective behaviours during a pandemic: A review. British Journal of Health Psychology, 15(4), 797–824. https://doi.org/10.1348/135910710X485826

Bults, M., Beaujean, D. J. M. A., De Zwart, O., Kok, G., Van Empelen, P., Van Steenbergen, J. E.,… Voeten, H. A. C. M. (2011). Perceived risk, anxiety, and behavioural responses of the general public during the early phase of the Influenza A (H1N1) pandemic in the Netherlands: Results of three consecutive online surveys. BMC Public Health, 11, 1–13. https://doi.org/10.1186/1471-2458-11-2

Bults, M., Beaujean, D. J. M. A., Richardus, J. H., & Voeten, H. A. C. M. (2015). Perceptions and behavioral responses of the general public during the 2009 influenza A (H1N1) pandemic: A systematic review. Disaster Medicine and Public Health Preparedness, 9(2), 207–219. https://doi.org/10.1017/dmp.2014.160

Centers for Disease Control and Prevention. (2020). Water and COVID-19 FAQs. Retrieved March 15, 2020, from https://www.cdc.gov/coronavirus/2019-ncov/php/water.html

Collinson, S., Khan, K., & Heffernan, J. M. (2015). The effects of media reports on disease spread and important public health measurements. PLoS ONE, 10(11), 1–21. https://doi.org/10.1371/journal.pone.0141423

Day, M. (2020). Covid-19: four fifths of cases are asymptomatic, China figures indicate. BMJ, 369, m1375. https://doi.org/10.1136/bmj.m1375

Diefenbach, M. A., & Leventhal, H. (1996). The common-sense model of illness representation: Theoretical and practical considerations. Journal of Social Distress and the Homeless, 5(1), 11–38. https://doi.org/10.1007/BF02090456

Ecker, U. K. H., Lewandowsky, S., Chang, E. P., & Pillai, R. (2014). The effects of subtle misinformation in news headlines. Journal of Experimental Psychology: Applied, 20(4), 323–335. https://doi.org/10.1037/xap0000028

Garfin, D. R., Silver, R. C., & Holman, E. A. (2020). The novel coronavirus (COVID-2019) outbreak: Amplification of public health consequences by media exposure. Health Psychology. https://doi.org/10.1037/hea0000875

Guan, W.-J., Ni, Z.-Y., Hu, Y., Liang, W.-H., Ou, C.-Q., He, J.-X., … China Medical Treatment Expert Group for Covid-19. (2020). Clinical Characteristics of Coronavirus Disease 2019 in China. The New England Journal of Medicine, 1–13. https://doi.org/10.1056/NEJMoa2002032

Han, P. K. J., Moser, R. P., & Klein, W. M. P. (2006). Perceived ambiguity about cancer prevention recommendations: Relationship to perceptions of cancer preventability, risk, and worry. Journal of Health Communication, 11(SUPPL. 1), 51–69. https://doi.org/10.1080/10810730600637541

Idler, E. L., & Benyamini, Y. (1997). Self-rated health and mortality: a review of twenty-seven community studies. Journal of Health and Social Behavior, 38(1), 21–37.

Johanson, S., Gross, S., & Cavataro, R. (2020, February 29). Chinese eateries fear closures. Sydney Morning Herald. Retrieved from https://search-proquest-com.www.proxy1.library.unsw.edu.au/docview/2366496854?accountid=12763

Klemm, C., Das, E., & Hartmann, T. (2016). Swine flu and hype: A systematic review of media dramatization of the H1N1 influenza pandemic. Journal of Risk Research, 19(1), 1–20. https://doi.org/10.1080/13669877.2014.923029

Lau, J. T. F., Kim, J. H., Tsui, H. Y., & Griffiths, S. (2007). Anticipated and current preventive behaviors in response to an anticipated human-to-human H5N1 epidemic in the Hong Kong Chinese general population. BMC Infectious Diseases, 7. https://doi.org/10.1186/1471-2334-1187-1118

Lauer, S. A., Grantz, K. H., Bi, Q., Jones, F. K., Zheng, Q., Meredith, H. R., … Lessler, J. (2020).The Incubation Period of Coronavirus Disease 2019 (COVID-19) From Publicly Reported Confirmed Cases: Estimation and Application. Annals of Internal Medicine, 2019. https://doi.org/10.7326/M20-0504

Minister for Health. (2020). First confirmed case of novel coronavirus in Australia. Retrieved from https://www.health.gov.au/ministers/the-hon-greg-hunt-mp/media/first-confirmed-case-of-novel-coronavirus-in-australia

Mizumoto, K., Kagaya, K., Zarebski, A., & Chowell, G. (2020). Estimating the asymptomatic proportion of coronavirus disease 2019 (COVID-19) cases on board the Diamond Princess cruise ship, Yokohama, Japan, 2020. Eurosurveillance, 25(10), 2000180. https://doi.org/10.2807/1560-7917.ES.2020.25.10.2000180

Nishiura, H., Kobayashi, T., Miyama, T., Suzuki, A., Jung, S., Hayashi, K., … Linton, N. M. (2020). Estimation of the asymptomatic ratio of novel coronavirus infections (COVID-19). MedRxiv, (February), 2020.02.03.20020248. https://doi.org/10.1101/2020.02.03.20020248

Paek, H.-J., & Hove, T. (2017). Risk Perceptions and Risk Characteristics. Oxford Research Encyclopedia of Communication, (March), 1–16. https://doi.org/10.1093/acrefore/9780190228613.013.283

Petrie, K., Faasse, K., & Thomas, M. G. (2016). Public Perceptions and Knowledge of the Ebola Virus, Willingness to Vaccinate, and Likely Behavioral Responses to an Outbreak. Disaster Medicine and Public Health Preparedness, 1–7. https://doi.org/10.1017/dmp.2016.67

Prime Minister of Australia. (2020). Media release: extention of travel ban to protect Australians from the coronavirus. Retrieved from https://www.pm.gov.au/media/extension-travel-ban-protect-australians-coronavirus

Rubin, G. J., Amlot, R., Page, L., & Wessely, S. (2009). Public perceptions, anxiety, and behaviour change in relation to the swine flu outbreak: cross sectional telephone survey. BMJ, 339, b2651. https://doi.org/10.1136/bmj.b2651

Simpson, C. R., Beever, D., Challen, K., De Angelis, D., Fragaszy, E., Goodacre, S., … Knight, M. (2019). The UK’s pandemic influenza research portfolio: a model for future research on emerging infections. The Lancet Infectious Diseases, 19(8), e295–e300. https://doi.org/10.1016/S1473-3099(18)30786-2

Slovic, P., Finucane, M. L., Peters, E., & MacGregor, D. G. (2007). The affect heuristic. European Journal of Operational Research, 177, 1333–1352. https://doi.org/10.1016/j.ejor.2005.04.006

Tang, C. S. K., & Wong, C.-Y. (2003). An outbreak of the severe acute respiratory syndrome: predictors of health behaviors and effect of community prevention measures in Hong Kong, China. American Journal of Public Health, 93(11), 1887–1888. https://doi.org/10.2105/AJPH.93.11.1887

Vosoughi, S., Roy, D., & Aral, S. (2018). The spread of true and false news online. Science, 359, 1146–1151.

Wang, C., Pan, R., Wan, X., Tan, Y., Xu, L., Ho, C. S., & Ho, R. C. (2020). Immediate psychological responses and associated factors during the initial stage of the 2019 coronavirus disease (COVID-19) epidemic among the general population in China. International Journal of Environmental Research and Public Health, 17(5). https://doi.org/10.3390/ijerph17051729

Weinstein, N D. (1988). The precaution adoption process. Health Psychology, 7(4), 355–386. Retrieved from http://ovidsp.ovid.com/ovidweb.cgi?T=JS&CSC=Y&NEWS=N&PAGE=fulltext&D=med3&AN=3049068

Weinstein, Neil D., Kwitel, A., McCaul, K. D., Magnan, R. E., Gerrard, M., & Gibbons, F. X. (2007). Risk perceptions: Assessment and relationship to influenza vaccination. Health Psychology, 26(2), 146–151. https://doi.org/10.1037/0278-6133.26.2.146

Wheaton, M. G., Abramowitz, J. S., Berman, N. C., Fabricant, L. E., & Olatunji, B. O. (2012). Psychological predictors of anxiety in response to the H1N1 (Swine Flu) Pandemic. Cognitive Therapy and Research, 36, 210–218.

Wikipedia. (2020). Misinformation related to the 2019–20 coronavirus outbreak.

World Health Organization. (2020a). Coronavirus disease (COVID-19) advice for the public: Myth busters. Retrieved March 8, 2020, from https://www.who.int/emergencies/diseases/novel-coronavirus-2019/advice-for-public/myth-busters

World Health Organization. (2020b). Modes of transmission of virus causing COVID-19: implications for IPC precaution recommendations. https://doi.org/10.1056/NEJMoa2001316.5.

World Health Organization. (2020c). Rolling updates on coronavirus disease (COVID-19). Retrieved March 7, 2020, from https://www.who.int/emergencies/diseases/novel-coronavirus-2019/events-as-they-happen

World Health Organization. (2020d). WHO Director-General’s opening remarks at the media briefing on COVID-19 - 3 March 2020. Retrieved March 21, 2020, from https://www.who.int/dg/speeches/detail/who-director-general-s-opening-remarks-at-the-media-briefing-on-covid-19---11-march-2020

Worldometer. (2020). Worldometer Coronavirus Australia. Retrieved April 7, 2020, from https://www.worldometers.info/coronavirus/country/australia/

Zimet, G. D., Rosberger, Z., Fisher, W. A., Perez, S., & Stupiansky, N. W. (2013). Beliefs, behaviors and HPV vaccine: Correcting the myths and the misinformation. Preventive Medicine, 57, 414–418. https://doi.org/10.1016/j.ypmed.2013.05.013

Zou, L., Ruan, F., Huang, M., Liang, L., Huang, H., Hong, Z., … Wu, J. (2020). SARS-CoV-2 Viral Load in Upper Respiratory Specimens of Infected Patients. The New England Journal of Medicine, 1, 19–21. https://doi.org/10.1056/NEJMc2000231

